# Sex-Specific Associations of the Plasma-Proteome with incident Coronary Artery Disease

**DOI:** 10.1101/2024.02.17.24302702

**Authors:** Vincent Q. Sier, Ko Willems van Dijk, Diana van Heemst, Paul H.A. Quax, J. Wouter Jukema, Raymond Noordam, Margreet R. de Vries

**Author notes:** shared last (Raymond Noordam, Margreet R. de Vries).

## Abstract

**Background and Aims:** The aetiology of coronary artery disease (CAD) is different for men and women, yet insights into underlying sex-specific biological and pathophysiological mechanisms are limited. We investigated the sex-specific associations of the plasma-proteome with incident CAD.

**Methods:** In 40,829 participants from UK Biobank free-of-CAD from baseline to 365 days thereafter (55% women, mean 56.9±8.1 years), we analysed associations between 2,922 plasma-proteins and CAD incidence. Baseline plasma samples (2006-2010), were analysed in relation to incident CAD over a median follow-up of 13.7 (IQR: 13.1,14.4) years. Combined and sex-specific analyses were performed using Cox-proportional hazard models, adjusting for considered confounders, and causal inference using Mendelian Randomisation (MR).

**Results:** Multivariable-adjusted Cox-proportional hazard models identified 1,138 proteins associated with incident CAD (false-discovery-rate-corrected p-value<0.05), of which 219 showed evidence for potential causality using MR. Overrepresentation analyses identified involvement of cytokine-cytokine receptor interactions (p<0.0001), matrix remodelling (p<0.0001), regulation of innate and adaptive immune cells (p<0.0001), and angiogenesis (p<0.0001) pathways associated with incident CAD. Sex-specific analyses revealed additional 412 female-exclusive and 37 male-exclusive proteins and distinct CAD-risk pathways were identified for women (e.g., innate immune response) and men (e.g., tube morphogenesis (angiogenesis)). Translation toward druggability on targets with causal evidence revealed sex-specific clinical drug candidates such as C1S (men) and FOXO1 (women).

**Conclusions:** Although the majority of proteins showed consistent associations with incident CAD in both sexes, multiple proteins and biological pathways were either more strongly associated with incident CAD in men or in women, potentially indicating sex-specific pathogenesis and opening new alleys for prevention and clinical strategies.

Visual abstract

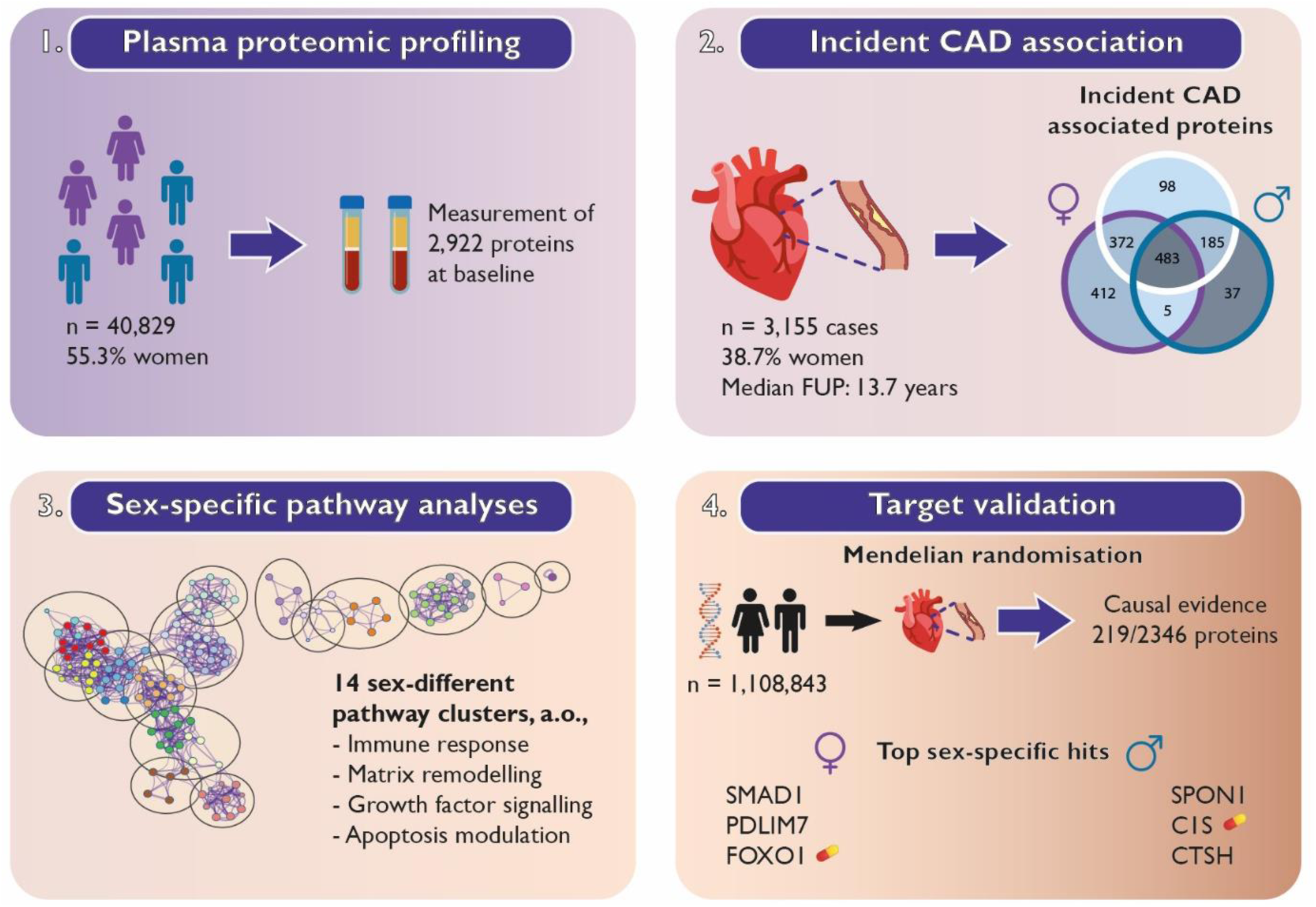

## 1. Introduction

Despite the growing recognition of disparities between men and women in the development, manifestation, and epidemiology of coronary artery disease (CAD),^1–5^ a critical gap persists in our understanding of its underlying sex-specific biological mechanisms. Both men and women are susceptible to develop ischaemic heart disease upon exposure to traditional risk factors.^6,7^ However, sex-specific aetiologies have been observed, with factors such as smoking, diabetes, and stress disproportionally affecting CAD incidence in women.^8–11^ Clinically, females present significantly more frequently with myocardial ischaemia with nonobstructive coronary arteries (INOCA), whereas males present more commonly with classical obstructive CAD.^12^ Regarding the latter, differences are observed in plaque composition, vulnerability, and burden between men and women, pointing toward a respectively more atheromatous versus fibrous phenotype.^13–15^

The plasma proteome reflects the dynamic biological state, which, prior to disease, may provide insight into the mechanisms of (sex-specific) CAD onset. The UK Biobank is a large-scale biomedical healthcare database comprising population-scale data of 2,922 unique plasma proteins in 54,219 individuals.^16,17^ Illustratively, using the UK Biobank data, a recent study demonstrated the significance of large-scale integration of plasma protein levels and genomics.^16^

Omics approaches, including genomics, transcriptomics, metabolomics, and proteomics, are powerful tools to unravel underlying biological mechanisms of disease, and may also provide insights into the sex-specific onset of CAD.^17–19^ A limited number of genomic studies has identified multiple sex-specific candidate genes as risk factors for CAD, within risk loci that encode for proteins that were primarily engaged in processes involved in lipid metabolism and vascular remodelling.^20–22^ Focusing on lesion development, human regulatory network analyses using patient material have indicated the existence of sex-specific plaque sub-phenotypes^13,23^ and cell signatures.^24,25^

Translating experimental findings into the identification of druggable targets is vital to enable clinical application. With the advent of increasingly complex datasets, advanced computational tools like interaction network analyses and artificial intelligence approaches facilitate the translation of novel targets to the clinic.^26,27^

We hypothesised that distinct proteomic patterns and pathways are associated with CAD onset in men and women. Therefore, in this study, by integrating both multivariable-adjusted regressions in combination with Mendelian Randomisation (MR) approaches,^28^ we aimed to investigate sex-specific plasma proteomic associations with incident CAD and identify potentially causal and druggable protein targets.

## 2. Methods

### 2.1 Multivariable-adjusted regression

#### Study setting (UK Biobank)

The UK Biobank is a general population cohort, in which approximately 500,000 participants were prospectively followed after recruitment between 2006 and 2010 (more information available via: https://www.ukbiobank.ac.uk/).^29^ Participants were between 40 and 70 years of age at enrolment and were recruited from the general population. Recruitment took place via invitation letters to eligible adults registered to the National Health Services (NHS) and living within 25 miles from one of the assessment centres. Written informed consent was retrieved from all participants and ethical approval of the study was given by the North-West Multicentre Ethics Committee. The present study was accepted and completed under project 56340.

#### Study population

The UK Biobank Pharma Plasma Proteome (UKB-PPP) project is a collaboration between thirteen biopharmaceutical companies and the UK Biobank, with the purpose of studying blood protein biomarkers in relation to disease onset. Baseline plasma samples from 54,219 individuals were selected randomly (n=46,595), or by consortium member pre-selection (n=6,376) and COVID-19 repeat imaging study (n=1,268) protocols. A total of 20 participants were pre-selected by the consortium members and participated in the COVID-19 repeat study. For the current study, we restricted the analysis to participants of the UK Biobank from European ancestry to minimise population stratification bias. Moreover, individuals with a history of CAD or a CAD incident within 365 days after baseline were excluded for all analyses to minimise influences caused by reverse causation, resulting in a total cohort of 40,829 participants.

#### Exposure

Proteomic profiling on the UKB-PPP samples was performed using the Olink Explore 3072, which processes 2,941 protein analytes, corresponding to 2,923 unique proteins from the Cardiometabolic, Cardiometabolic_II, Inflammation, Inflammation_II, Neurology, Neurology_II, Oncology, and Oncology_II panels. Further details on the Olink explore platform and assays are available in the summary publication of the UKB-PPP.^16^ The current analyses have been performed in accordance with the quality consideration as described in Sun et al., excluding one protein (GLIPR1), due to >80% of data failing quality control (99.4%).^16^

#### Cardiovascular disease outcomes

A UK Biobank algorithm was employed to collect participant data on incident CAD, using data from the general practitioner, linked hospital admissions, death registries, and self-report. As coded according to the International Classification of Diseases (ICD) standards, the study outcome of CAD was defined as angina pectoris (I20), myocardial infarction (I21, I22), and acute and chronic ischaemic heart disease (I24, I25), whichever came first. Participants were followed until the occurrence of the CAD event, death, loss to follow-up or the end of follow-up, whichever came first.

#### Other variables

Data on age, sex, body mass index (BMI), insulin use, smoking, menopausal status (for women only), and use of cholesterol lowering medication were collected at the assessment centre of the UK Biobank through touchscreen questionnaire and physical measures. For smoking status, we included previous and current smokers. The Towsend Deprivation Index (TDI) was included as a marker for socio-economic status, and reflects the neighbourhood of the individual participant based on zip code. Non-insulin dependent type diabetes mellitus was obtained via linkage to health-related records.

#### Multivariable-adjusted Cox-proportional hazard models

All the statistics were performed in R version 3.6.1 statistical software (The R Foundation for Statistical Computing, Vienna, Austria). Characteristics of the study population were studied at baseline as means (with standard deviations), medians (with interquartile ranges; for non-normally distributed continuous variables), or proportions (for categorical variables only).

Cox-proportional hazard models were performed to investigate the associations between standardised plasma protein levels (in standard deviation increase in level) and incident CAD in participants free-of-CAD at baseline or within 365 days thereafter (Survival package in R^30^). In these analyses, participants were observed until the conclusion of the follow-up period, loss to follow-up or mortality, whichever came first. Regression analyses were adjusted for baseline age, sex, BMI, TDI, baseline diagnosis of diabetes mellitus, insulin use, smoking, and use of cholesterol-lowering medication.

Analyses were performed for the total study population, as well as stratified for men and women. Additional female analyses were performed to adjust and additionally restrict for post-menopausal status. To provide evidence favouring heterogeneity by sex, we additionally included a multiplicative interaction term between the protein level and sex in the analyses on incident CAD, adjusted for the confounders. We corrected for multiple testing using the false-discovery-rate (FDR).

For the analysis of enriched biological pathways in our data, we used the integrated biological database Metascape (web-based, more information available via: http://metascape.org).^31^ In short, Metascape facilitates the translation of large gene sets to involvement in biological processes via computed overrepresentation. More specifically, Metascape incorporates multiple ontologies for enrichment analysis and eliminates redundancies following hierarchical clustering of terms based on enrichment. The platform leverages >40 independent knowledgebases, among which the ontology resources Molecular Signatures Database (MSigDB), Kyoto Encyclopedia of Genes and Genomes (KEGG), Gene Ontology (GO) knowledgebase, Reactome Pathway Database, and WikiPathways.^32–36^

### 2.2 Mendelian randomisation

To provide evidence favouring possible causal associations, we performed two-sample MR analyses. For the single-nucleotide polymorphisms (SNP)-exposure associations, we used the data from the SNPs associated with the protein levels from UK Biobank.^16^ From these, we selected all independent lead SNPs associated with protein levels (both cis and trans SNPs) with a SNP-exposure p-value <1.7e-11, considering multiple testing for the number of proteins in the genome-wide association study. As SNP-outcome datasets, we used summary-level genome-wide association data from the Coronary ARtery DIsease Genome wide Replication and Meta-analysis plus the Coronary Artery Disease Genetics consortium (CARDIoGramplusC4D) (60,801 cases and 123,504 controls),^37^ UK Biobank (122,733 cases and 424,528 controls), and freeze 9 from the Finngen biobank (https://www.finngen.fi/en) (43,518 cases and 333,759 controls), resulting in a total of 227,052 cases and 881,791 controls. In each dataset, CAD was defined as angina pectoris (I20), myocardial infarction (I21, I22), and acute and chronic ischaemic heart disease (I24, I25). With the exception of the CARDIoGRAMplusC4D dataset which comprised 23% of non-European ancestry participants, all other datasets only used participants from European ancestry. The studies contributing data to these (meta-)analyses have been approved by the necessary local medical ethics committees and all participants contributing to these efforts provided written informed consent. For the purpose of the present study, we only used the summary-level data and no individual-level data. Data were only available for all participants combined and not stratified by sex.

#### Mendelian Randomisation meta-analysis

The primary analysis for MR employed inverse-variance weighted (IVW) regression analysis, assuming the absence of invalid genetic instruments such as directional pleiotropy.^38^ All MR analyses utilised the R-based package “TwoSample MR”. The mean effect estimate was derived separately from each outcome database through fixed-effect IVW meta-analysis of Wald ratios (gene-outcome [log odds ratio] divided by gene-exposure associations) estimated for each instrumental variable.^39^ Results were expressed as odds ratios (ORs) for CAD risk. Under the assumption of met MR criteria, this odds ratio served as an estimate of the causal effect of the exposure on the outcome.

To assess potential violations of main MR assumptions stemming from directional pleiotropy, MR-Egger regression analyses and weighted-median estimator analyses were performed.^39–41^ The MR-Egger’s intercept estimated the average pleiotropic effect across genetic variants, with a (significant) non-zero value indicating bias in the IVW estimate.^40^ The weighted-median estimator yielded a consistent valid estimate if at least half of the instrumental variables were valid.^41^ All analyses were performed separately for the individual GWAS summary datasets, and subsequently meta-analysed using fixed-effects with the R-based “rmeta” package.

### 2.3 Druggability analyses

To evaluate target druggability, we used the machine learning platform DrugnomeAI to calculate aggregated probability scores based on a combined clinically-approved (Tclin) and clinical-phase (Tier 1) drug candidate model.^42^ In short, DrugnomeAI is an adaptation of mantis-ml, a machine-learning framework, based on a stochastic semi-supervised learning approach.^42,43^

Essentially, DrugnomeAI employs a process in which the machine learning algorithm learns from both labelled and unlabelled data in a probabilistic manner. Multiple resources have been integrated in the assessment of target druggability within DrugnomeAI, among which databases on protein-protein, drug-gene, and chemical-gene interactions such as Pharos, Drug Gene Interaction Database (DGIdb), and Comparative Toxicogenomics Database (CTDbase).^44–46^

## 3. Results

### 3.1 Primary analysis UK Biobank

#### 3.1.1 Summary information cases and controls

From the UKB-PPP project cohort, a total of 40,829 individuals were included free-of-CAD at baseline and in the first year after inclusion, with a mean age of 56.9 (standard deviation 8.10) years, 55.3% women, and a mean BMI of 27.0 kg/m^2^ (Table 1). Cholesterol-lowering and insulin medication use at recruitment were 14.6% and 0.9% respectively, with 7.3% of total participants being diagnosed with non-insulin dependent diabetes mellitus. Moreover, there was a mean TDI (socio-economic status score) of –1.52 (3.01) and 44.6% of the individuals were past and/or current smoker (Table 1).

**Table 1:** Baseline characteristics of the study population.

|  | Baseline (n=40,829) | Women | Men |
| --- | --- | --- | --- |
| **Sex** |  |  |  |
| Female (%) | 22,590 (55.3%) | 22,590 (100%) |  |
| Male (%) | 18,239 (44.7%) |  | 18,239 (100%) |
| **Age at recruitment (years)** |  |  |  |
| Mean (SD) | 56.9 (8.10) | 56.8 (8.01) | 57.0 (8.22) |
| **Body mass index** |  |  |  |
| Mean (SD) | 27.3 (4.71) | 27.0 (5.08) | 27.7 (4.16) |
| **Cholesterol-lowering medication at recruitment** |  |  |  |
| Yes (%) | 5,975 (14.6%) | 2,512 (11.1%) | 3,463 (19.0%) |
| **Insulin use at recruitment** |  |  |  |
| Yes (%) | 374 (0.9%) | 166 (0.7%) | 208 (1.1%) |
| **Smoking** |  |  |  |
| Past & current (%) | 18,202 (44.6%) | 9,101 (40.3%) | 9,101 (49.9%) |
| **Townsend deprivation index (socio-economic)** |  |  |  |
| Mean (SD) | -1.52 (3.01) | -1.54 (2.95) | -1.56 (2.90) |
| **Non-insulin dependent diabetes mellitus** |  |  |  |
| Yes (%) | 2,996 (7.3%) | 1,249 (5.5%) | 1,747 (9.6%) |

#### 3.1.2 Proteomic associations with CAD

After a median follow-up time of 13.7 (IQR: 13.1, 14.4) years, there were 3,155 incident CAD cases, of which 38.7% women. In combined sex multivariate analysis, 1,138 out of 2,922 proteins were significantly associated with incident CAD after correction for multiple comparisons (FDR) and adjusting for considered confounding factors (Figure 1a, Supplementary Table 1). Separate sex-specific analyses revealed 412 female– and 37 male-specific CAD proteins (Figure 1b, Supplementary Figure 1, Supplementary Table 2, Supplementary Table 3). Adjustment nor restriction of women for post-menopausal status did substantively change the results (Supplementary Figure 2, Supplementary Table 4).

**Figure 1.**
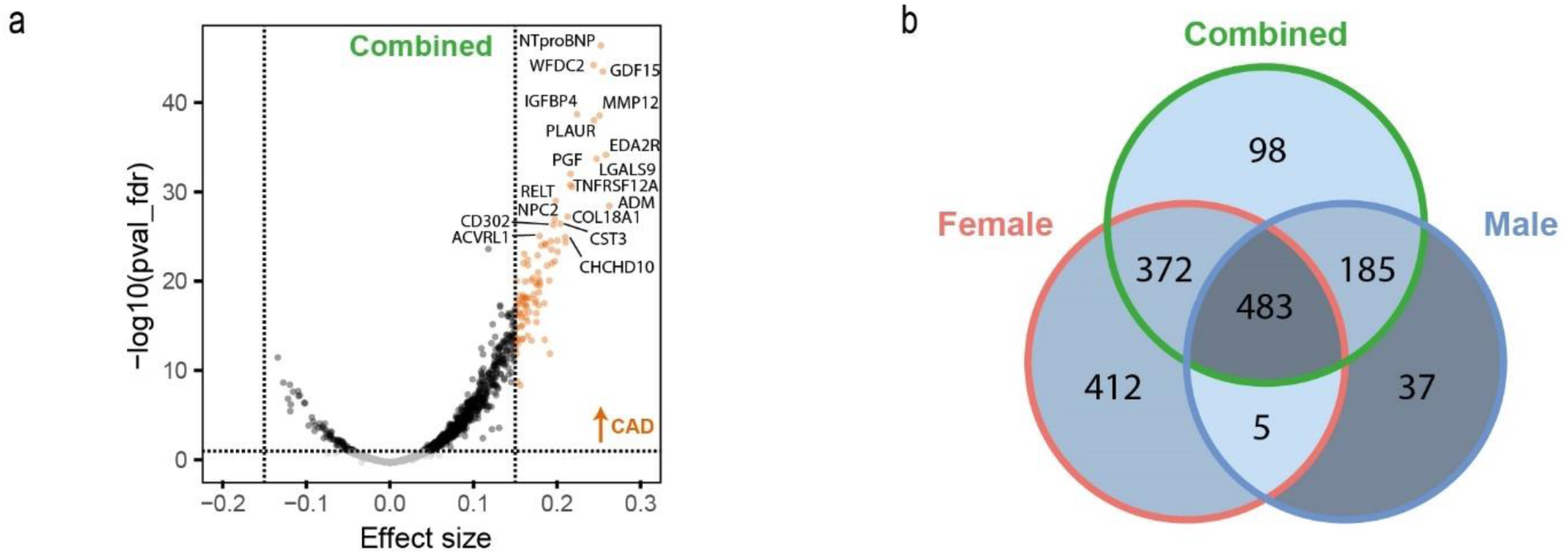
Proteomic associations for incident CAD. (**a**) Volcano plot representing the effect size of combined-sex plasma-proteomic profiles for CAD (FDR<0.05, abs(β) ≥ 0.15). Orange dots denote positive significant associations with incident CAD. The proteins were part of the Cardiometabolic, Cardiometabolic_II, Inflammation, Inflammation_II, Neurology, Neurology_II, Oncology, and Oncology_II panels of the Olink Explore 3072 platform (**b**) Venn diagram of proteins associated with CAD for the sex-combined (green), female-specific (red), and male-specific (blue) Cox-proportional hazard models. FDR: false discovery rate

For graphical reasons only, we filtered for effect sizes ≥0.15 across the combined, female-, and male-specific analyses, resulting in 158 proteins of interest associated with incident CAD. Considering this cut-off, this resulted in 57 combined, 67 female-specific, and 37 male-specific proteins for visualisation (Figure 2).

**Figure 2.**
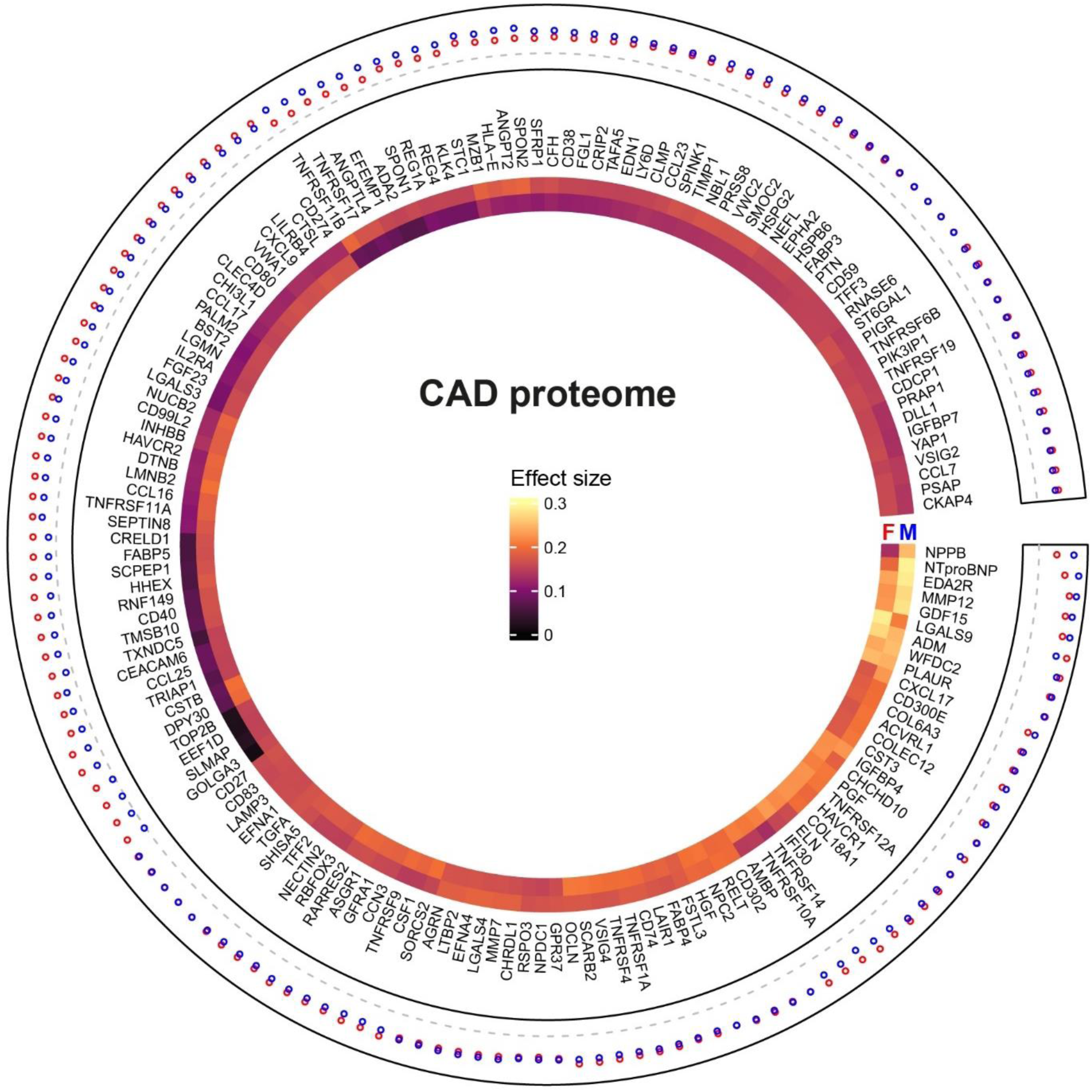
Circular heatmap for proteins associated with increased hazard for incident CAD for men and women. Average effect sizes per protein are represented by the heatmap layer women (red; inner) and men (blue; outer) respectively. General clustering was plotted in a counter-clockwise fashion around the heatmap, based on increasing effect sizes and computed sex differences.

#### 3.1.3 Sex-specific proteomic pathways

Based on the complete CAD-incidence-associated plasma-proteome in women and men, we performed pathway analyses. This revealed sex-specific differences associated with incident CAD (Figure 3, Supplementary Table 5, Supplementary Table 6). Notable distinctions were observed between both sexes, including angiogenic (tube morphogenesis) and hormonal response (Insulin-like growth factor response) components in men, and the innate immune response and programmed cell death regulation in women (Figure 3a, 3b). Moreover, we also found multiple shared pathways between both sexes, among which those corresponding to the matrisome, cytokine-cytokine receptor interactions, regulation of cell migration, neutrophil degranulation, and enzyme-linked receptor protein signalling.

**Figure 3.**
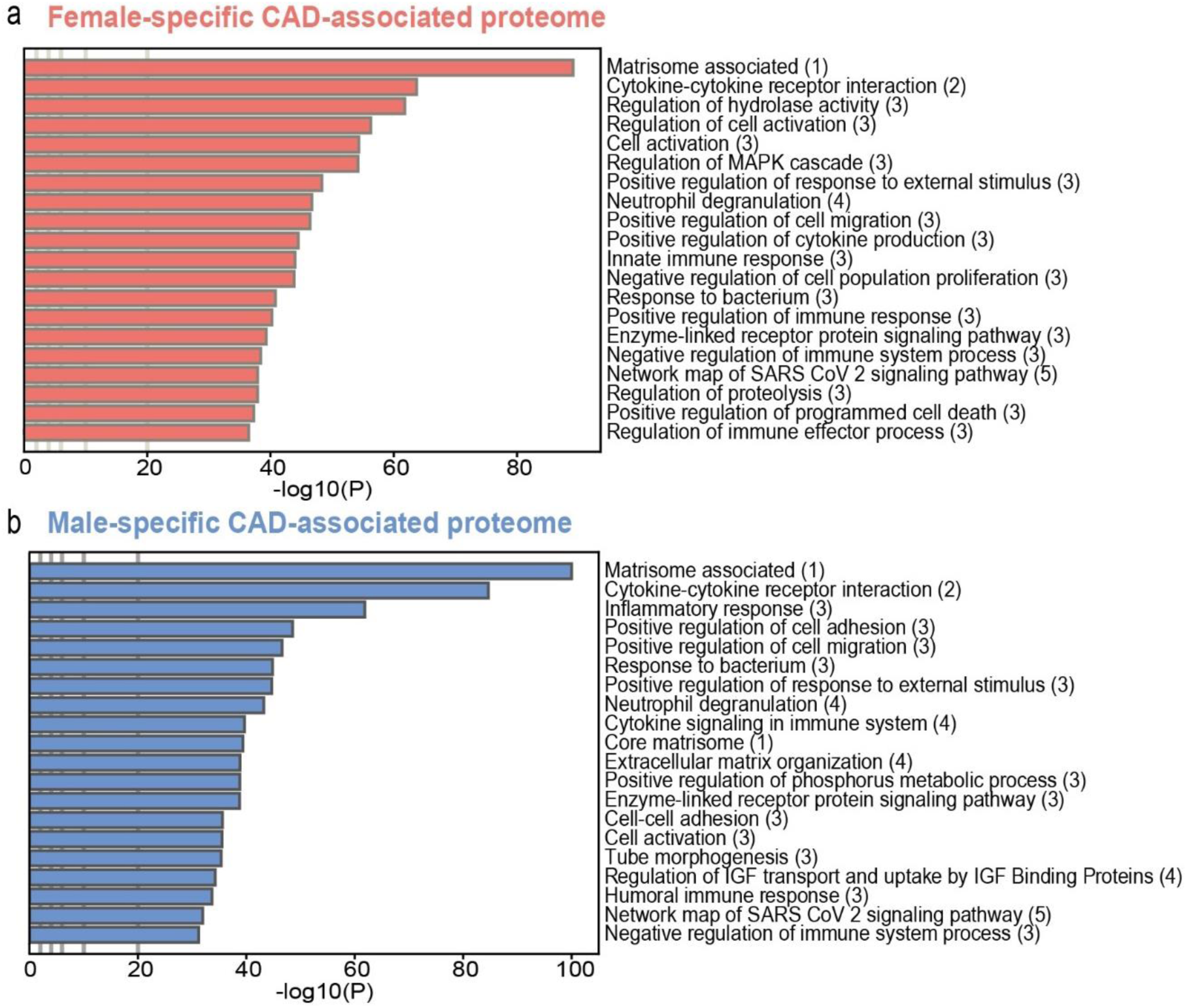
Sex-specific proteome pathway analyses for CAD incidence. (**a, b**) Pathway analyses of respectively the female and male CAD-associated proteome. Multiple ontology sources were included to cluster all enriched terms into groups. *(1)MSigDB: Molecular Signatures Database; (2)KEGG: Kyoto Encyclopedia of Genes and Genomes; (3)GO: Gene Ontology knowledgebase;(4)Reactome; (5)WP: WikiPathways*.

#### 3.1.4 Sex-different protein interaction analyses

Based on interaction analyses (male-versus-female),^47^ we found effect sizes of 711 proteins to be significantly different between men and women (FDR<0.05; Figure 4a, Supplementary Table 7). Based on these significantly different effect sizes in men versus women, we performed pathway analyses and subsequent network auto-clustering, and identified fourteen independent pathway clusters (Figure 4b, Supplementary Table 8). In general, the fourteen clusters represented differences between the CAD-associated proteome of men and women pertaining to innate and adaptive immunity, matrix remodelling, growth factor responses, apoptosis modulation, and lipid management. The three largest clusters were “response innate immune”, ‘pathogenic infection virus”, and “cysteine endopeptidase regulation” (Figure 4b).

**Figure 4.**
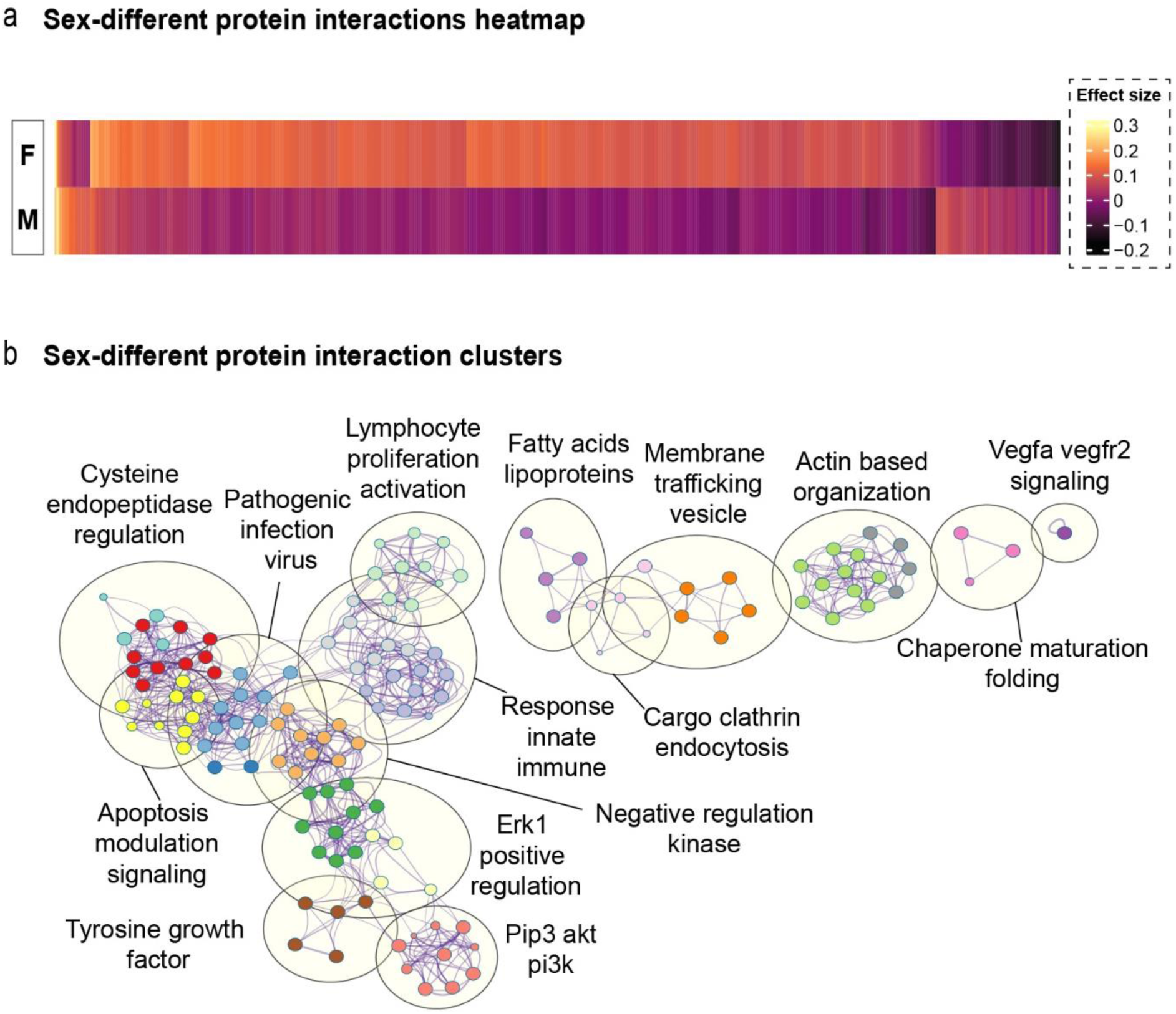
Sex-different protein interactions. (**a**) Sex-protein interaction heatmap of significantly different proteins associated with CAD onset between men and women (n=711) (**b**) Network auto-clustering based on significant interaction clusters for incident CAD. Nodes represent individual pathways corresponding to the higher order cluster.

### 3.2 MR meta-analysis

We performed two-sample MR analyses on 2,346 proteins for which there were genetic instruments,^28^ and observed evidence for possible positive causal associations for 219 proteins in the meta-analysis of the three individual GWAS, of which 66 proteins had a logodds per s.d. higher level than 0.15 (Figure 5a, Supplementary Table 9).

**Figure 5.**
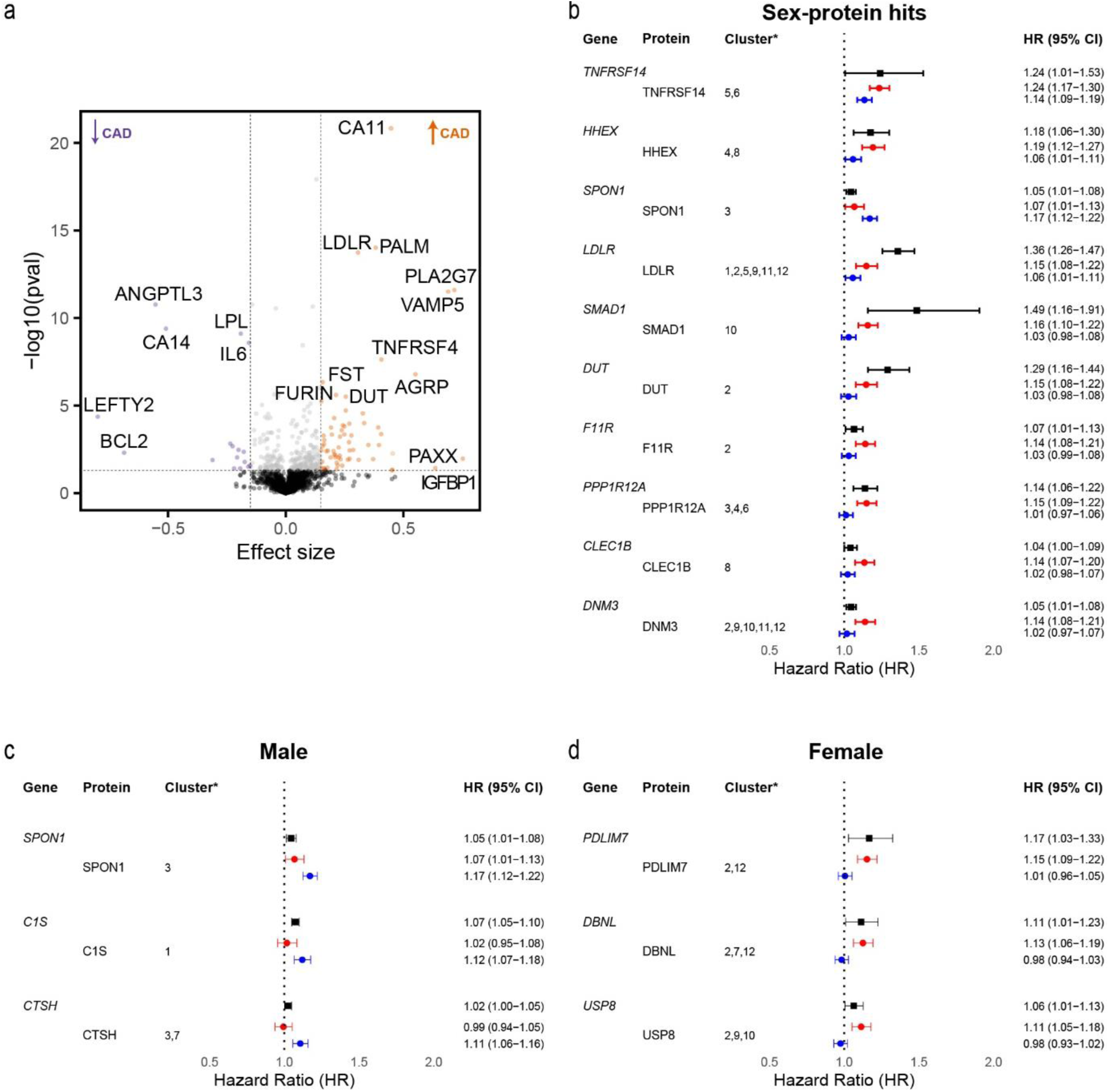
Causal evidence for sex-specific target associations with CAD. (**a**) Volcano plot of genetic MR associations with CAD in the combined UK Biobank-FinnGen-CARDIoGRAMplusC4D cohort. Orange and purple dots denote significant positive and negative associations respectively (FDR<0.05, abs(β) ≥ 0.15). (**b**) Forest plot depicting the top 10 target protein hazard ratios resulting from the sex-protein interaction analyses, as causally validated by the MR meta-analysis data (black) and jointly sorted for women (red) and men (blue). (**c, d**) Forest plot depicting the top three protein hazard ratios for CAD, as sorted for male– (**c**) and female-specific (**d**) differential top hits. *Clusters according to the interaction-analyses: *1: response innate immune, 2: actin based organisation, 3: cysteine endopeptidase regulation, 4: negative regulation kinase, 5: pathogenic infection virus, 6: erk1 positive regulation, 7: apoptosis modulation signalling, 8: lymphocyte proliferation activation, 9: membrane trafficking vesicle, 10: tyrosine growth factor, 11: cargo clathrin endocytosis, 12: fatty acids lipoprotein, 13: chaperone maturation folding, 14: vegfa vegfr2 signalling*.

To identify the sex-specific proteins with the highest hazard for CAD incidence, we analysed the 219 proteins for occurrence within our fourteen sex-protein interaction clusters (Supplementary Table 10). In total, we identified 59 hits that pertained to varying groups of clusters, of which the ten with the highest combined hazard ratio are shown in Figure 5b.

When considering the highest absolute differences between both sexes, Spondin 1 (SPON1), Complement C1s (C1S) and Cathepsin H (CTSH) emerged as additional top hits for men, while PDZ And LIM Domain 7 (PDLIM7), Drebrin like (DBNL), and Ubiquitin specific peptidase (USP8) were prominent within the specific top hits for women. Further examination revealed sex-specific nuances of the data within the framework of the fourteen pathway clusters (Figure 4b, Supplementary Table 10).

### 3.3 Top target druggability

Based on the 219 significant targets found through the MR analyses, we identified the top 20 hits associated with CAD across both sexes for the combined average of female and male hazard ratios as shown in Table 2. To assess druggability of these targets, we used aggregated probability scores of our top 20 hits based on a clinically-approved and clinical-phase drug candidate model (Table 2, Supplementary Table 11).

**Table 2:**
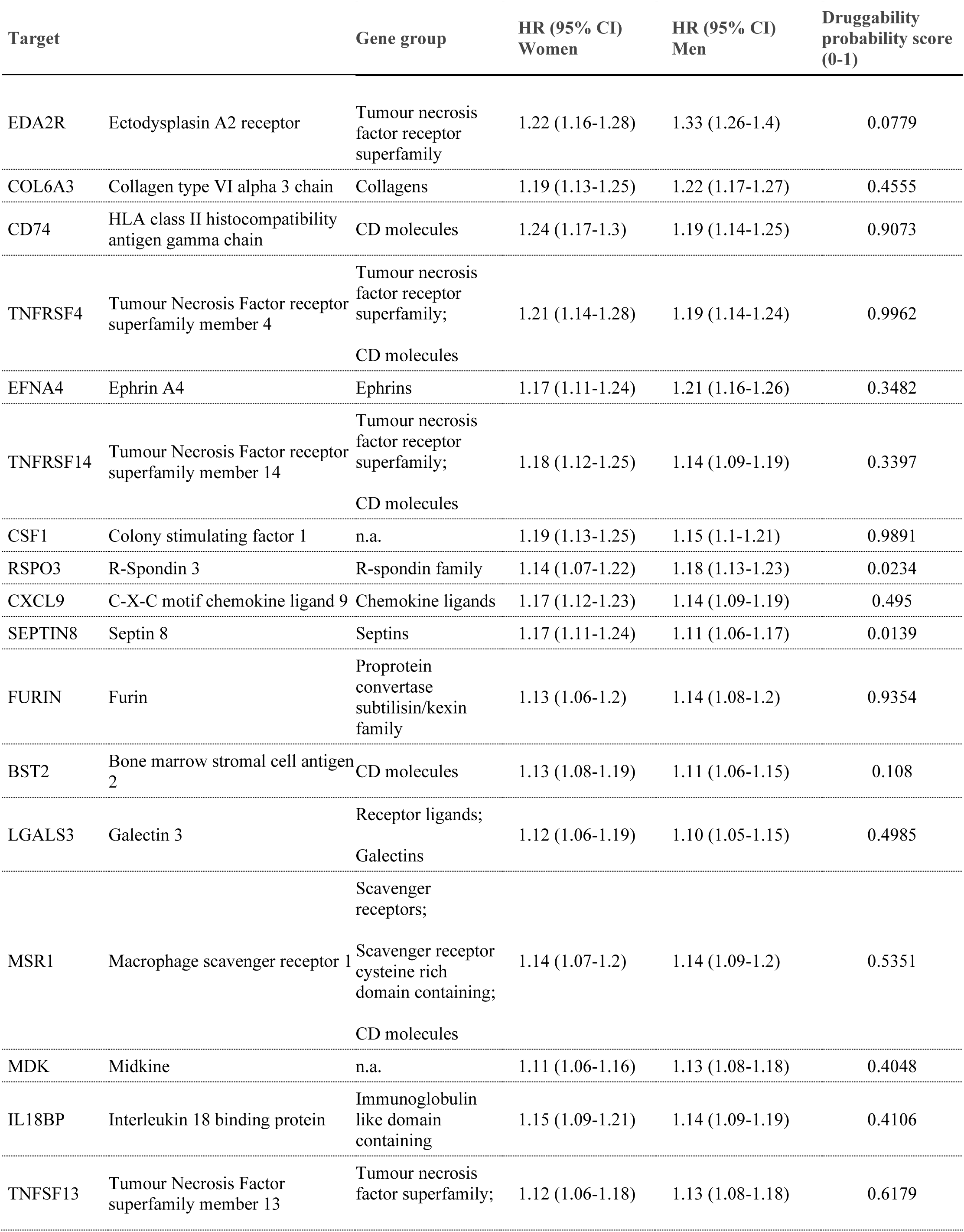
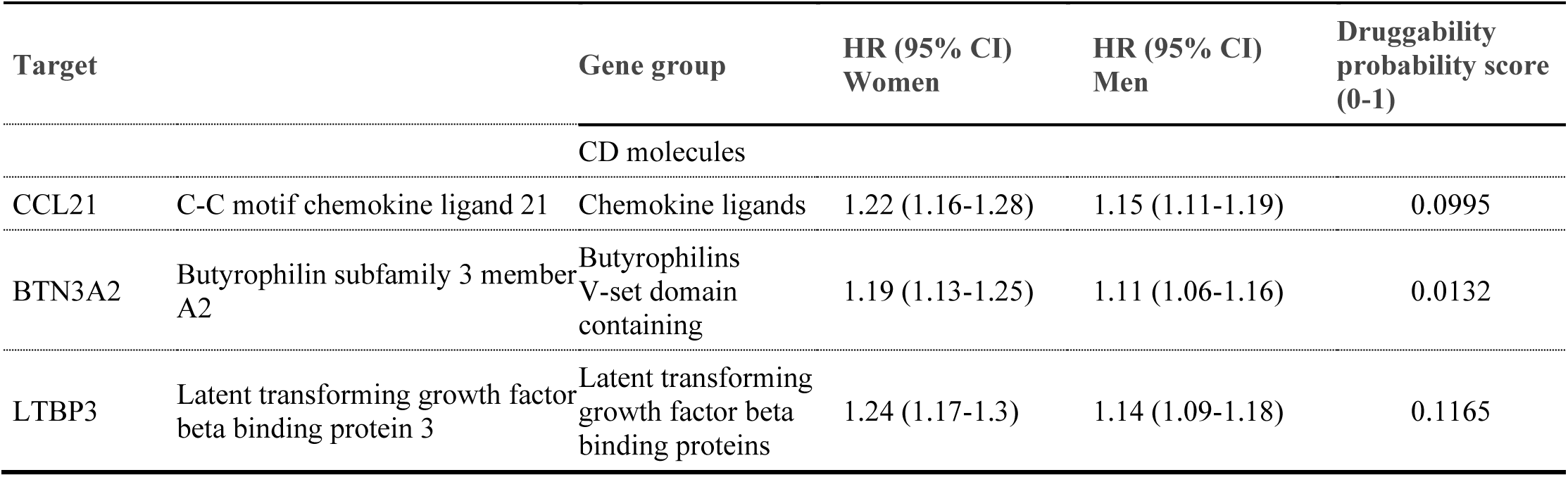
Druggability of top 20 combined targets.

From this integrated data model, five targets emerged with high druggability probability scores surpassing the threshold of 0.5. Notably, TNFRSF4, CSF1, FURIN, CD74, TNFSF13 demonstrated potential for therapeutic intervention. (Table 2). Further analyses were performed using the sex-specific targets as identified for men and women. Within the previously identified male-specific top hits, the complement system activation component C1S had the highest druggability score (0.8578), followed by CTSH (0.5118) (Table 3). In the context of the female top hits, the first target with high druggability was Forkhead box protein 1 (FOXO1, 0.6286) (Supplementary Table 11).

**Table 3:** Druggability of top three female– and male-specific proteins.

| Target |  | Gene group | HR (95% CI)<br>Women | HR (95% CI)<br>Men | Druggability<br>probability<br>score (0-1) |
| --- | --- | --- | --- | --- | --- |
| *Women* |  |  |  |  |  |
| PDLIM7 | PDZ and LIM<br>domain 7 | PDZ domain<br>containing<br>LIM domain<br>containing | 1.01 (0.96-1.05) | 1.15 (1.09-1.22) | 0.05630 |
| DBNL | Drebrin like | MicroRNA protein<br>coding host genes | 0.96 (0.94-1.03) | 1.13 (1.06-1.19) | 0.1764 |
| USP8 | Ubiquitin specific<br>peptidase 8 | Ubiquitin specific<br>peptidases | 0.96 (0.93-1.02) | 1.11 (1.05-1.18) | 0.04340 |
| *Men* |  |  |  |  |  |
| SPON1 | Spondin 1 | n.a. | 1.17 (1.12-1.22) | 1.07 (1.01-1.13) | 0.09470 |
| C1S | Complement C1S | Sushi domain<br>containing<br>Complement system<br>activation components | 1.12 (1.07-1.18) | 1.02 (0.95-1.08) | 0.8578 |
| CTSH | Cathepsin H | Cathepsins<br>Minor<br>histocompatibility<br>antigens | 1.11 (1.06-1.16) | 0.99 (0.94-1.05) | 0.5118 |

## 4. Discussion

This study observed the presence of sex-specific differences in the plasma proteome associated with incident CAD. Through integrated genomic and proteomic approaches, we observed differences between men and women in pathways related to matrix organisation, cytokine interactions, regulation of innate and adaptive immune cells, angiogenesis, and growth factor signalling related to incident CAD. Furthermore, we provided evidence favouring causal relations in a sex-combined study sample. These findings granted new insights into sex-specific pathophysiological mechanisms and implications for incident CAD, and allowed for the identification of druggable targets unique to men and women.

Despite apparent differences in top target findings of previous plasma proteomic approaches,^16,17,48–50^ we found evidence for the contribution of shared biological pathways leading to CAD. Notably, from our protein-sex interaction analyses it appeared that specific pathways, such as vascular endothelial growth factor (VEGF) signalling, are associated with sex differences in CAD development. Previously, our group and others have shown that dysfunctional VEGF signalling is involved in leaky plaque neovessels and can subsequently contribute to lesion destabilisation.^51–53^ Non-mature neovessels promote plaque development as the endothelium functions as a significant entrance route for immune cells via NF-κB promotion and subsequent TNF signalling.^54,55^ Endothelial dysfunction has been described in both sexes within the context of atherosclerotic disease. Men are more likely to suffer from intraplaque haemorrhage secondary to neovessel-rupture, whereas women more often present with coronary microvascular dysfunction (CMD) and subsequent angina with non-obstructive coronary arteries.^56–58^ This is noteworthy, considering that female hormone signalling has been associated with CMD,^59^ while oestrogen replacement therapy has not been found effective to halt classical obstructive atherosclerotic plaque progression in post-menopausal women with ischaemic heart disease.^60,61^ The findings of our present study further substantiate the concept that in the development of CAD, both sexes exhibit numerous common risk factors in addition to sex-specific mechanisms. Specifically, our results highlight the involvement of various conventional protein risk factors such as the well-described targets LDLR and PCSK9, which are shared between women and men.

After integration of large-scale plasma proteomics with MR analyses, we found evidence of high druggability for 5/20 of our strongest CAD-associated hits (TNFRSF4, TNFSF13, CSF1, FURIN, CD74). Indeed, tumour necrosis factor levels have previously been associated with higher risk of coronary artery disease and specific drugs targeting TNFRSF4 and TNFSF13 have been employed in clinical trials in patients with respectively T– and B-cell related auto-immune diseases such as IGA nephropathy (Sibeprenlimab) and atopic dermatitis (telazorlimab).^62–66^ CSF1 is a known driver of atherosclerosis through monocyte/macrophage activation, proliferation, and differentiation.^67^ High levels of CSF1 expression in the tumour micro-environment have been related to poor prognosis in solid tumours, in which context numerous monoclonal antibody therapies have reached clinical trials with mixed results.^68^ Recently, FURIN, a member of the proprotein convertases, was found by Mazidi et al. as their most strongly associated protein with ischaemic heart disease^17^ and was previously linked to atherogenesis via upregulation of TNFSF13.^69–71^ Another hit, CD74, is expressed on monocytes, macrophages, and B-cells and is involved in antigen presentation. Direct inhibition of CD74 through the monoclonal antibody milatuzumab has been described in haematologic malignancies.^72,73^ Another interesting approach is to induce CD74 degradation through inhibition of the cysteine protease Cathepsin S. Elevated serum cathepsin S levels are associated with plaque instability and vulnerability and a small molecule inhibitor has been described for patients with Sjögren’s syndrome (petesicatib) (clinicaltrials.gov: nct02701985). Taking sex-specificity into consideration, we identified 59 sex-specific associations with evidence for possible causality, of which C1S (complement factor) and FOXO1 (transcription factor) were the top druggable hits in men and women respectively. From the literature, it is known that general C1 inhibition diminishes early intimal hyperplasia. Conversely, deficiencies in C1 inhibitory protein function are associated with hereditary angioedema and subsequent microvascular endothelial dysfunction within the atherosclerotic plaque, drawing parallels with the tube morphogenesis pathway in our male-specific analyses.^74–76^ Although we have found many more proteins to be specifically associated with incident CAD in women compared to men, despite the lower number of cases in women, the strongest protein-CAD associations in women did not seem to yield strong evidence as druggable targets using currently available databases. The first druggable target in women was FOXO1, a transcription factor which has also been described to be involved in the pathogenesis of polycystic ovary syndrome and glucose metabolism.^77,78^ Altogether, our findings point toward sex-different pathways and factors in CAD development, and suggest sex-specific drug candidates among our identified hits.

### Strengths & limitations

The primary strength of our study is the large sample size and in particular the proportion of included women, enabling the analysis of a comprehensive pool of plasma proteins in association with CAD while correcting for multiple testing and confounders. The integration of genomics and proteomics provided evidence for causality. Limitations include the focus on individuals of European ancestry, and therefore these results apply to only that part of the population. We did not have access to sex-stratified summary-level datasets of GWAS on CAD. For this reason, we were not able to perform MR analyses in men and women separately. Alternatively, we performed MR analyses in a large sample of men and women combined. Furthermore, in the multivariable-regression analyses, we excluded participants who developed CAD within the first year of follow-up to limit the potential influence of reverse causation. Nevertheless, this approach does not limit the potential presence of sex-specific residual confounding, although adjustment nor restriction of women for postmenopausal status did materially differ the results, suggesting such confounding might be limited.

## Conclusion

In summary, by linking the sex-specific plasma-proteome to CAD incidence and providing evidence for causal relations, we were able to identify strongly-associated female– and male-specific proteins along with their biological pathways. These findings offer potential directions for developing targeted preventive and interventional strategies against CAD.

## Data Availability

The present study has been conducted using the UK Biobank Resource (Application Number 56,340) that is available to researchers. Summary data are available from the original sources. Summary-level data used for the two-sample Mendelian Randomisation analyses are available online through the infrastructure of the R package twosampleMR or on the website of the FinnGen Biobank (https://www.finngen.fi/en)

## Author contribution statement

Conceptualisation; VQS, JWJ, PHAQ, RN, MRdV. Methodology; VQS, RN. Formal analysis; VQS, RN. Resources; RN, KWvD, DvH. Writing-Original Draft; VQS, RN, MRdV. Writing-Review & Editing; VQS, KWvD, DvH, JWJ, PHAQ, RN, MRdV. Visualisation; VQS. Supervision; PHAQ, RN, MRdV, Project Administration; RN, MRdV. Funding acquisition; VQS, PHAQ, RN, MRdV.

## References

1. Crea F, Battipaglia I, Andreotti F. Sex differences in mechanisms, presentation and management of ischaemic heart disease. Atherosclerosis 2015;241:157–168. doi: 10.1016/j.atherosclerosis.2015.04.802

2. Reynolds HR, Shaw LJ, Min JK, et al. Association of Sex With Severity of Coronary Artery Disease, Ischemia, and Symptom Burden in Patients With Moderate or Severe Ischemia: Secondary Analysis of the ISCHEMIA Randomized Clinical Trial. JAMA Cardiol 2020;5:773–786. doi: 10.1001/jamacardio.2020.0822

3. Aggarwal NR, Patel HN, Mehta LS, et al. Sex Differences in Ischemic Heart Disease: Advances, Obstacles, and Next Steps. Circ Cardiovasc Qual Outcomes 2018;11:e004437. doi: 10.1161/CIRCOUTCOMES.117.004437

4. Galbraith M, Drossart I. Disparity in diagnosis and treatment of cardiovascular disease in women: a call to joint action from the European Society of Cardiology Patient Forum. Eur Heart J 2024;45:79–80. doi: 10.1093/eurheartj/ehad480

5. Leening MJ, Ferket BS, Steyerberg EW, et al. Sex differences in lifetime risk and first manifestation of cardiovascular disease: prospective population based cohort study. BMJ 2014;349:g5992. doi: 10.1136/bmj.g5992

6. Khot UN, Khot MB, Bajzer CT, et al. Prevalence of conventional risk factors in patients with coronary heart disease. JAMA 2003;290:898–904. doi: 10.1001/jama.290.7.898

7. Bello N, Mosca L. Epidemiology of coronary heart disease in women. Prog Cardiovasc Dis 2004;46:287–295. doi: 10.1016/j.pcad.2003.08.001

8. Mathur P, Ostadal B, Romeo F, Mehta JL. Gender-Related Differences in Atherosclerosis. Cardiovasc Drugs Ther 2015;29:319–327. doi: 10.1007/s10557-015-6596-3

9. Parvand M, Rayner-Hartley E, Sedlak T. Recent Developments in Sex-Related Differences in Presentation, Prognosis, and Management of Coronary Artery Disease. Can J Cardiol 2018;34:390–399. doi: 10.1016/j.cjca.2018.01.007

10. Peters SA, Huxley RR, Woodward M. Diabetes as risk factor for incident coronary heart disease in women compared with men: a systematic review and meta-analysis of 64 cohorts including 858,507 individuals and 28,203 coronary events. Diabetologia 2014;57:1542–1551. doi: 10.1007/s00125-014-3260-6

11. Manfrini O, Yoon J, van der Schaar M, et al. Sex Differences in Modifiable Risk Factors and Severity of Coronary Artery Disease. J Am Heart Assoc 2020;9:e017235. doi: 10.1161/JAHA.120.017235

12. Bairey Merz CN, Pepine CJ, Walsh MN, Fleg JL. Ischemia and No Obstructive Coronary Artery Disease (INOCA): Developing Evidence-Based Therapies and Research Agenda for the Next Decade. Circulation 2017;135:1075–1092. doi: 10.1161/CIRCULATIONAHA.116.024534

13. Sakkers TR, Mokry M, Civelek M, et al. Sex differences in the genetic and molecular mechanisms of coronary artery disease. Atherosclerosis 2023;384:117279. doi: 10.1016/j.atherosclerosis.2023.117279

14. Seegers LM, Araki M, Nakajima A, et al. Sex Differences in Culprit Plaque Characteristics Among Different Age Groups in Patients With Acute Coronary Syndromes. Circ Cardiovasc Interv 2022;15:e011612. doi: 10.1161/CIRCINTERVENTIONS.121.011612

15. Mautner SL, Lin F, Mautner GC, Roberts WC. Comparison in women versus men of composition of atherosclerotic plaques in native coronary arteries and in saphenous veins used as aortocoronary conduits. J Am Coll Cardiol 1993;21:1312–1318. doi: 10.1016/0735-1097(93)90302-h

16. Sun BB, Chiou J, Traylor M, et al. Plasma proteomic associations with genetics and health in the UK Biobank. Nature 2023;622:329–338. doi: 10.1038/s41586-023-06592-6

17. Mazidi M, Wright N, Yao P, et al. Plasma Proteomics to Identify Drug Targets for Ischemic Heart Disease. J Am Coll Cardiol 2023;82:1906–1920. doi: 10.1016/j.jacc.2023.09.804

18. van der Harst P, Verweij N. Identification of 64 Novel Genetic Loci Provides an Expanded View on the Genetic Architecture of Coronary Artery Disease. Circ Res 2018;122:433–443. doi: 10.1161/CIRCRESAHA.117.312086

19. McGarrah RW, Crown SB, Zhang GF, Shah SH, Newgard CB. Cardiovascular Metabolomics. Circ Res 2018;122:1238–1258. doi: 10.1161/CIRCRESAHA.117.311002

20. Liu LY, Schaub MA, Sirota M, Butte AJ. Sex differences in disease risk from reported genome-wide association study findings. Hum Genet 2012;131:353–364. doi: 10.1007/s00439-011-1081-y

21. Consortium CAD, Deloukas P, Kanoni S, et al. Large-scale association analysis identifies new risk loci for coronary artery disease. Nat Genet 2013;45:25–33. doi: 10.1038/ng.2480

22. Huang Y, Hui Q, Gwinn M, et al. Sexual Differences in Genetic Predisposition of Coronary Artery Disease. Circ Genom Precis Med 2021;14:e003147. doi: 10.1161/CIRCGEN.120.003147

23. Hartman RJG, Owsiany K, Ma L, et al. Sex-Stratified Gene Regulatory Networks Reveal Female Key Driver Genes of Atherosclerosis Involved in Smooth Muscle Cell Phenotype Switching. Circulation 2021;143:713–726. doi: 10.1161/CIRCULATIONAHA.120.051231

24. Lu C, Donners M, Karel J, et al. Sex-specific differences in cytokine signaling pathways in circulating monocytes of cardiovascular disease patients. Atherosclerosis 2023;384:117123. doi: 10.1016/j.atherosclerosis.2023.04.005

25. Talukdar HA, Foroughi Asl H, Jain RK, et al. Cross-Tissue Regulatory Gene Networks in Coronary Artery Disease. Cell Syst 2016;2:196–208. doi: 10.1016/j.cels.2016.02.002

26. Qureshi R, Irfan M, Gondal TM, et al. AI in drug discovery and its clinical relevance. Heliyon 2023;9:e17575. doi: 10.1016/j.heliyon.2023.e17575

27. Dara S, Dhamercherla S, Jadav SS, Babu CM, Ahsan MJ. Machine Learning in Drug Discovery: A Review. Artif Intell Rev 2022;55:1947–1999. doi: 10.1007/s10462-021-10058-4

28. Lawlor DA, Tilling K, Davey Smith G. Triangulation in aetiological epidemiology. Int J Epidemiol 2016;45:1866–1886. doi: 10.1093/ije/dyw314

29. Sudlow C, Gallacher J, Allen N, et al. UK biobank: an open access resource for identifying the causes of a wide range of complex diseases of middle and old age. PLoS Med 2015;12:e1001779. doi: 10.1371/journal.pmed.1001779

30. Therneau TM, Grambsch PM. Modeling Survival Data: Extending the Cox Model: Springer New York; 2013.

31. Zhou Y, Zhou B, Pache L, et al. Metascape provides a biologist-oriented resource for the analysis of systems-level datasets. Nat Commun 2019;10:1523. doi: 10.1038/s41467-019-09234-6

32. Liberzon A, Birger C, Thorvaldsdottir H, et al. The Molecular Signatures Database (MSigDB) hallmark gene set collection. Cell Syst 2015;1:417–425. doi: 10.1016/j.cels.2015.12.004

33. Kanehisa M, Goto S. KEGG: kyoto encyclopedia of genes and genomes. Nucleic Acids Res 2000;28:27–30. doi: 10.1093/nar/28.1.27

34. Harris MA, Clark J, Ireland A, et al. The Gene Ontology (GO) database and informatics resource. Nucleic Acids Res 2004;32:D258–261. doi: 10.1093/nar/gkh036

35. Milacic M, Beavers D, Conley P, et al. The Reactome Pathway Knowledgebase 2024. Nucleic Acids Res 2024;52:D672–D678. doi: 10.1093/nar/gkad1025

36. Slenter DN, Kutmon M, Hanspers K, et al. WikiPathways: a multifaceted pathway database bridging metabolomics to other omics research. Nucleic Acids Res 2018;46:D661–D667. doi: 10.1093/nar/gkx1064

37. Nikpay M, Goel A, Won HH, et al. A comprehensive 1,000 Genomes-based genome-wide association meta-analysis of coronary artery disease. Nat Genet 2015;47:1121–1130. doi: 10.1038/ng.3396

38. Burgess S, Butterworth A, Thompson SG. Mendelian randomization analysis with multiple genetic variants using summarized data. Genet Epidemiol 2013;37:658–665. doi: 10.1002/gepi.21758

39. Staley JR, Burgess S. Semiparametric methods for estimation of a nonlinear exposure-outcome relationship using instrumental variables with application to Mendelian randomization. Genet Epidemiol 2017;41:341–352. doi: 10.1002/gepi.22041

40. Bowden J, Davey Smith G, Burgess S. Mendelian randomization with invalid instruments: effect estimation and bias detection through Egger regression. Int J Epidemiol 2015;44:512–525. doi: 10.1093/ije/dyv080

41. Bowden J, Davey Smith G, Haycock PC, Burgess S. Consistent Estimation in Mendelian Randomization with Some Invalid Instruments Using a Weighted Median Estimator. Genet Epidemiol 2016;40:304–314. doi: 10.1002/gepi.21965

42. Raies A, Tulodziecka E, Stainer J, et al. DrugnomeAI is an ensemble machine-learning framework for predicting druggability of candidate drug targets. Commun Biol 2022;5:1291. doi: 10.1038/s42003-022-04245-4

43. Vitsios D, Petrovski S. Mantis-ml: Disease-Agnostic Gene Prioritization from High-Throughput Genomic Screens by Stochastic Semi-supervised Learning. Am J Hum Genet 2020;106:659–678. doi: 10.1016/j.ajhg.2020.03.012

44. Sheils TK, Mathias SL, Kelleher KJ, et al. TCRD and Pharos 2021: mining the human proteome for disease biology. Nucleic Acids Res 2021;49:D1334–D1346. doi: 10.1093/nar/gkaa993

45. Freshour SL, Kiwala S, Cotto KC, et al. Integration of the Drug-Gene Interaction Database (DGIdb 4.0) with open crowdsource efforts. Nucleic Acids Res 2021;49:D1144–D1151. doi: 10.1093/nar/gkaa1084

46. Davis AP, Wiegers TC, Johnson RJ, et al. Comparative Toxicogenomics Database (CTD): update 2023. Nucleic Acids Res 2023;51:D1257–D1262. doi: 10.1093/nar/gkac833

47. Altman DG, Bland JM. Interaction revisited: the difference between two estimates. BMJ 2003;326:219. doi: 10.1136/bmj.326.7382.219

48. Williams SA, Ostroff R, Hinterberg MA, et al. A proteomic surrogate for cardiovascular outcomes that is sensitive to multiple mechanisms of change in risk. Sci Transl Med 2022;14:eabj9625. doi: 10.1126/scitranslmed.abj9625

49. Valdes-Marquez E, Clarke R, Hill M, Watkins H, Hopewell JC. Proteomic profiling identifies novel independent relationships between inflammatory proteins and myocardial infarction. Eur J Prev Cardiol 2023;30:583–591. doi: 10.1093/eurjpc/zwad020

50. Bom MJ, Levin E, Driessen RS, et al. Predictive value of targeted proteomics for coronary plaque morphology in patients with suspected coronary artery disease. EBioMedicine 2019;39:109–117. doi: 10.1016/j.ebiom.2018.12.033

51. de Vries MR, Parma L, Peters HAB, et al. Blockade of vascular endothelial growth factor receptor 2 inhibits intraplaque haemorrhage by normalization of plaque neovessels. J Intern Med 2019;285:59–74. doi: 10.1111/joim.12821

52. Sluiter TJ, Tillie R, de Jong A, et al. Myeloid PHD2 Conditional Knockout Improves Intraplaque Angiogenesis and Vascular Remodeling in a Murine Model of Venous Bypass Grafting. J Am Heart Assoc 2024:e033109. doi: 10.1161/JAHA.123.033109

53. Dunmore BJ, McCarthy MJ, Naylor AR, Brindle NP. Carotid plaque instability and ischemic symptoms are linked to immaturity of microvessels within plaques. J Vasc Surg 2007;45:155–159. doi: 10.1016/j.jvs.2006.08.072

54. Parma L, Baganha F, Quax PHA, de Vries MR. Plaque angiogenesis and intraplaque hemorrhage in atherosclerosis. Eur J Pharmacol 2017;816:107–115. doi: 10.1016/j.ejphar.2017.04.028

55. Sluiter TJ, van Buul JD, Huveneers S, Quax PHA, de Vries MR. Endothelial Barrier Function and Leukocyte Transmigration in Atherosclerosis. Biomedicines 2021;9. doi: 10.3390/biomedicines9040328

56. Sidik NP, Stanley B, Sykes R, et al. Invasive Endotyping in Patients With Angina and No Obstructive Coronary Artery Disease: A Randomized Controlled Trial. Circulation 2024;149:7–23. doi: 10.1161/CIRCULATIONAHA.123.064751

57. Reynolds HR, Diaz A, Cyr DD, et al. Ischemia With Nonobstructive Coronary Arteries: Insights From the ISCHEMIA Trial. JACC Cardiovasc Imaging 2023;16:63–74. doi: 10.1016/j.jcmg.2022.06.015

58. van Dam-Nolen DHK, van Egmond NCM, Dilba K, et al. Sex Differences in Plaque Composition and Morphology Among Symptomatic Patients With Mild-to-Moderate Carotid Artery Stenosis. Stroke 2022;53:370–378. doi: 10.1161/STROKEAHA.121.036564

59. Tunc E, Eve AA, Madak-Erdogan Z. Coronary Microvascular Dysfunction and Estrogen Receptor Signaling. Trends Endocrinol Metab 2020;31:228–238. doi: 10.1016/j.tem.2019.11.001

60. Hodis HN, Mack WJ, Azen SP, et al. Hormone therapy and the progression of coronary-artery atherosclerosis in postmenopausal women. N Engl J Med 2003;349:535–545. doi: 10.1056/NEJMoa030830

61. Herrington DM, Reboussin DM, Brosnihan KB, et al. Effects of estrogen replacement on the progression of coronary-artery atherosclerosis. N Engl J Med 2000;343:522–529. doi: 10.1056/NEJM200008243430801

62. Yuan S, Carter P, Bruzelius M, et al. Effects of tumour necrosis factor on cardiovascular disease and cancer: A two-sample Mendelian randomization study. EBioMedicine 2020;59:102956. doi: 10.1016/j.ebiom.2020.102956

63. Mathur M, Barratt J, Chacko B, et al. A Phase 2 Trial of Sibeprenlimab in Patients with IgA Nephropathy. N Engl J Med 2024;390:20–31. doi: 10.1056/NEJMoa2305635

64. Rewerska B, Sher LD, Alpizar S, et al. Phase 2b randomized trial of OX40 inhibitor telazorlimab for moderate-to-severe atopic dermatitis. J Allergy Clin Immunol Glob 2024;3:100195. doi: 10.1016/j.jacig.2023.100195

65. Monraats PS, Pires NM, Schepers A, et al. Tumor necrosis factor-alpha plays an important role in restenosis development. Faseb j 2005;19:1998–2004. doi: 10.1096/fj.05-4634com

66. Monraats PS, Pires NM, Agema WR, et al. Genetic inflammatory factors predict restenosis after percutaneous coronary interventions. Circulation 2005;112:2417–2425. doi: 10.1161/CIRCULATIONAHA.105.536268

67. Sinha SK, Miikeda A, Fouladian Z, et al. Local M-CSF (Macrophage Colony-Stimulating Factor) Expression Regulates Macrophage Proliferation and Apoptosis in Atherosclerosis. Arterioscler Thromb Vasc Biol 2021;41:220–233. doi: 10.1161/ATVBAHA.120.315255

68. Cannarile MA, Weisser M, Jacob W, et al. Colony-stimulating factor 1 receptor (CSF1R) inhibitors in cancer therapy. J Immunother Cancer 2017;5:53. doi: 10.1186/s40425-017-0257-y

69. Turpeinen H, Raitoharju E, Oksanen A, et al. Proprotein convertases in human atherosclerotic plaques: the overexpression of FURIN and its substrate cytokines BAFF and APRIL. Atherosclerosis 2011;219:799–806. doi: 10.1016/j.atherosclerosis.2011.08.011

70. Yakala GK, Cabrera-Fuentes HA, Crespo-Avilan GE, et al. FURIN Inhibition Reduces Vascular Remodeling and Atherosclerotic Lesion Progression in Mice. Arterioscler Thromb Vasc Biol 2019;39:387–401. doi: 10.1161/ATVBAHA.118.311903

71. Sluijter JP, Verloop RE, Pulskens WP, et al. Involvement of furin-like proprotein convertases in the arterial response to injury. Cardiovasc Res 2005;68:136–143. doi: 10.1016/j.cardiores.2005.05.016

72. Martin P, Furman RR, Rutherford S, et al. Phase I study of the anti-CD74 monoclonal antibody milatuzumab (hLL1) in patients with previously treated B-cell lymphomas. Leuk Lymphoma 2015;56:3065–3070. doi: 10.3109/10428194.2015.1028052

73. Kaufman JL, Niesvizky R, Stadtmauer EA, et al. Phase I, multicentre, dose-escalation trial of monotherapy with milatuzumab (humanized anti-CD74 monoclonal antibody) in relapsed or refractory multiple myeloma. Br J Haematol 2013;163:478–486. doi: 10.1111/bjh.12565

74. Krijnen PA, Kupreishvili K, de Vries MR, et al. C1-esterase inhibitor protects against early vein graft remodeling under arterial blood pressure. Atherosclerosis 2012;220:86–92. doi: 10.1016/j.atherosclerosis.2011.10.021

75. Shagdarsuren E, Bidzhekov K, Djalali-Talab Y, et al. C1-esterase inhibitor protects against neointima formation after arterial injury in atherosclerosis-prone mice. Circulation 2008;117:70–78. doi: 10.1161/CIRCULATIONAHA.107.715649

76. Demirturk M, Polat N, Guz G, et al. There is an increased risk of atherosclerosis in hereditary angioedema. Int Immunopharmacol 2012;12:212–216. doi: 10.1016/j.intimp.2011.11.013

77. Xu R, Wang Z. Involvement of Transcription Factor FoxO1 in the Pathogenesis of Polycystic Ovary Syndrome. Front Physiol 2021;12:649295. doi: 10.3389/fphys.2021.649295

78. Lee S, Dong HH. FoxO integration of insulin signaling with glucose and lipid metabolism. J Endocrinol 2017;233:R67–R79. doi: 10.1530/JOE-17-0002

