## Supplementary Figures for "Sex-Specific Associations of the Plasma-Proteome with incident Coronary Artery Disease"

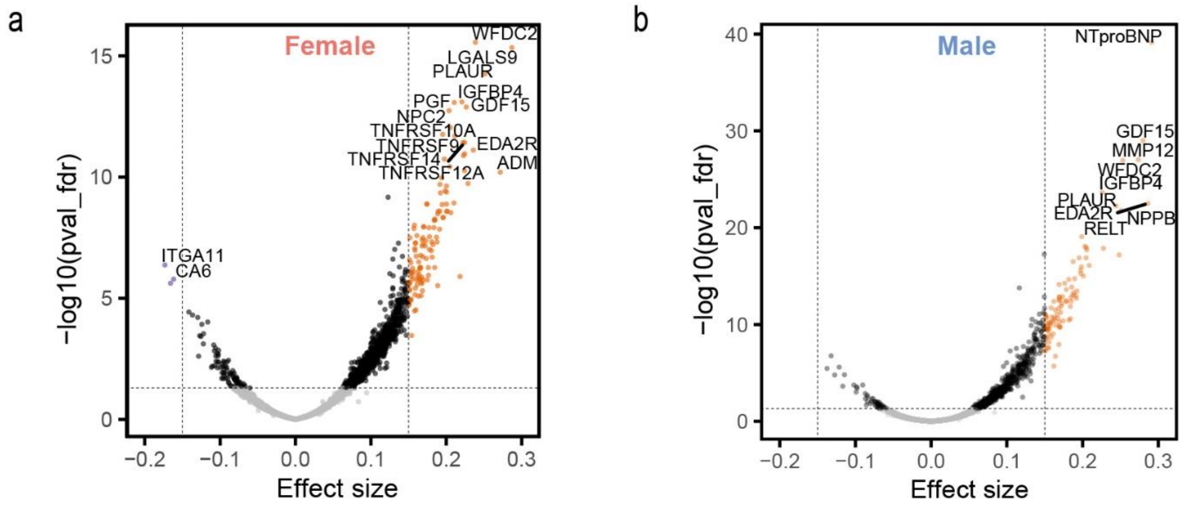


**Supplementary Figure 1**. **Sex-specific proteomic associations for incident CAD.** (**a**) Female- and (b) male-specific volcano plots representing the effect size of combined-sex plasma-proteomic profiles for CAD (FDR<0.05, abs(β) ≥ 0.15). Orange and purple dots respectively denote positive and negative significant associations with incident CAD. The proteins were part of the Cardiometabolic, Cardiometabolic_II, Inflammation, Inflammation_II, Neurology, Neurology_II, Oncology, and Oncology_II panels of the Olink Explore 3072 platform

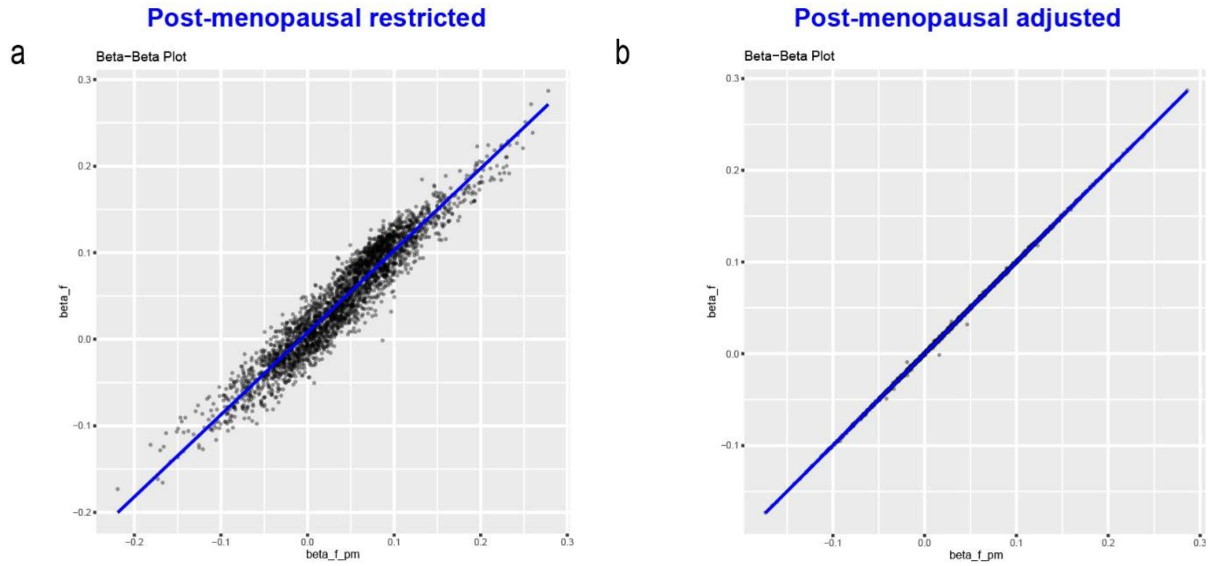


**Supplementary Figure 2**. **Post-menopausal analyses for protein associations with CAD.** Beta-beta plots for restriction (**a**) and (**b**) adjustment of women for postmenopausal status, relative to the non-adapted female-specific model.
